# SARS-CoV-2 serological tests can generate false positive results for samples from patients with chronic inflammatory diseases

**DOI:** 10.1101/2020.11.13.20231076

**Authors:** Nastya Kharlamova, Nicky Dunn, Sahl K Bedri, Svante Jerling, Malin Almgren, Francesca Faustini, Iva Gunnarsson, Johan Rönnelid, Rille Pullerits, Inger Gjertsson, Karin Lundberg, Anna Månberg, Elisa Pin, Peter Nilsson, Sophia Hober, Katharina Fink, Anna Fogdell-Hahn

## Abstract

**Objectives:** Patients with chronic inflammatory diseases are often treated with immunosuppressants and therefore are of particular concern during the SARS-CoV-2 pandemic. Serological tests will improve our understanding of the infection and immunity in this population, unless the tests give false positive results. The aim of this study was to evaluate the specificity of SARS-Cov-2 serological assays with samples from patients with chronic inflammatory diseases collected before April 2019, thus defined as negative.

**Methods:** Samples from patients with multiple sclerosis (MS, n=10), rheumatoid arthritis (RA, n=47) with or without rheumatoid factor (RF) and/or anti-cyclic citrullinated peptide antibodies (anti-CCP2) and RF +/- systemic lupus erythematosus (SLE, n=10), were tested with 17 commercially available lateral flow assays (LFA), two ELISA kits and one in-house developed multiplex bead-based assay.

**Results:** Six LFA and the in-house IgG assay gave the correct negative results for all samples. However, the majority of assays (n=13), gave false positive signal with samples from patients with RA and SLE. This was most notable in RF positive RA samples. MS samples did not give any false positive in any of the assays.

**Conclusion:** The majority of the verified serological assays were sensitive to interfering antibodies in samples from patients with chronic inflammatory diseases and therefore may have poor specificity in this context. For these patients, the risk of false positivity should be considered when interpreting results of the SARS-CoV-2 serological assays.

## Introduction

Severe acute respiratory syndrome coronavirus-2 (SARS-CoV-2) is an enveloped positive sense single stranded RNA virus and the causative agent of the coronavirus disease 2019 (COVID-19) which emerged as a pandemic in the human population late 2019.^1^ The cumulative number of infected and fatal cases can be followed at the Johns Hopkins University COVID-19 Dashboard.^2^ Patients with chronic inflammatory disease are often treated with immunomodulatory treatments and therefore potentially more susceptible to infections.^3^ As a result, there has been substantial concern during the pandemic as to the potential increased risk COVID-19 disease severity and mortality among these patient groups. To date, there is still limited evidence about their risk of severe COVID-19, or knowledge of how their disease or immunomodulatory treatment may affect either their pre-existing immunity or ability to develop protective immunity following infection.^4 5^

Approximately 6% of the world’s population are affected by chronic inflammatory diseases which includes conditions such as multiple sclerosis (MS), rheumatoid arthritis (RA) and systemic lupus erythematosus (SLE).^6^ These are generally progressive diseases and although for the majority there are no cures, treatment is centred around slowing disease progression with immunomodulatory treatments. The hallmarks of autoimmune diseases are inflammation, loss of self-tolerance and the presence of autoantibodies. MS is a chronic inflammatory disorder restricted to the central nervous system, characterized by demyelination, axonal loss and the formation of sclerotic plaques. The worldwide prevalence is estimated to be 2.2 million cases, but with large geographical variation.^7^ RA is a heterogeneous chronic inflammatory disease, which affected close to 5 million people globally by 2010 and with prevalence increasing due to the increased aging of the human population.^8^ The disease is characterized by synovial inflammation and the formation of the pannus which causes cartilage and bone destruction, joint dysfunction, pain and disability. Rheumatoid factor (RF) and anti-citrullinated protein antibodies (ACPA) are the most frequent and the most studied RA-related autoantibodies. RF is an antibody reactive with the Fc portion of IgG, mainly consisting of IgM in Caucasian RA populations, but also IgG and IgA RF are present. Although RF is detected in approximately 70% of RA patients, the presence of RF is not specific for RA. These autoantibodies are also present in a variety of other diseases as well as in the general population and may increase with age, smoking and chronic infection.^9 10^ SLE is a systemic inflammatory disease of the connective tissue, characterized by a loss of self-tolerance and leading to production and deposition of a large panel of autoantibodies and immune complexes formation.^11^ Clinical manifestation of SLE is heterogeneous and can affect multiple organs. Approximately 25% of SLE patients have RF.^12^

Serological tests are useful for determining past infection and present immunity. The presence of IgM antibodies indicates a recent infection, whereas presence of IgG antibodies indicates possible long-lasting immunity.^13^ Important information can be achieved by having access to reliable serological methods during a pandemic; to identify seropositive people for convalescent plasma donations; guide policies and ease restrictions on human mobility based on sero-epidemiological evidences; ensure immunity to allow key workers to return to work after exposure; and evaluate vaccine development studies and vaccine strategies.

Due to the substantial global demand, SARS-CoV-2 serological testing has been rapidly developed and released to the market. The assays are validated before release and also often independently verified before approved.^14 15^ However, the panel of samples used to determine specificity is often focused on ruling out cross-reactivity with other viral infections and seldom includes serum from patients with chronic inflammatory diseases.^15^ Based on experience from development and validation of serology assays for measuring anti-drug antibodies (ADA) in persons with chronic inflammatory disease, it is recommended to show specificity against drug naïve patient serum, as antibodies present in patients with autoimmune diseases are known to interact with reagents in serological assays and give unspecific signals.^16 17^ Given the significant role serological tests may have as useful large screening tools for immunity as the pandemic unfolds, it is important to verify the specificity of SARS-CoV-2 serological tests also for specific patient groups.

The aim of this study was to verify the specificity of commercially available SARS-CoV-2 serological tests, using a cohort of patients with different chronic inflammatory diseases with samples collected before the SARS-CoV-2 outbreak, as negative controls.

## Material and Methods

### Study design and ethical approval

This retrospective cohort study was approved by the Ethics Review Authority and all participants provided informed consent at the time of sample collection to participate in future ethically approved studies.

## Materials

### Patient serum samples

To evaluate specificity of SARS-CoV-2 serological assays in patients with chronic inflammatory diseases, a selection of negative control samples were retrieved from the biobank (n=68). To exclude individuals with risk of previous exposure to SARS-CoV-2 infection, only samples collected before April 2019 were included in the study. Serum samples were selected from patients with MS (n=10), RA (n=47, of which 2 samples were from the same patient), or SLE (n=10) (Table 1). MS patient samples were collected in a research laboratory doing routine testing for anti-drug antibodies (ADAs) at The Centre for Molecular Medicine, Stockholm and had been treated with interferon beta (IFNβ). Three MS samples were ADA positive. Of the RA samples, 40 were from the Swedish population-based case control study Epidemiological Investigation of RA (EIRA) and had not been treated with any disease modifying anti-rheumatic drug (DMARD).^18^ Of these patients, 20 were RF and anti-CCP2 positive (50%); six were RF negative but anti-CCP2 positive (15%), and 14 were both RF and anti-CCP2 negative (35%).^19^ The additional seven RA patient samples were retrieved from a prospective study cohort (Sahlgrenska University Hospital, Gothenburg) and were infliximab (IFX) treated. Of these seven patient samples, three were RF and anti-CCP2 positive; two were RF negative but anti-CCP2 positive; one was RF positive but anti-CCP2 negative, and one sample was both RF and anti-CCP2 negative. The SLE samples were obtained from a study investigating the development of ADA against rituximab (RTX), and therefore all patients were RTX treated. Only one of the ten samples was ADA positive (supplementary table 1).

**Table 1.**
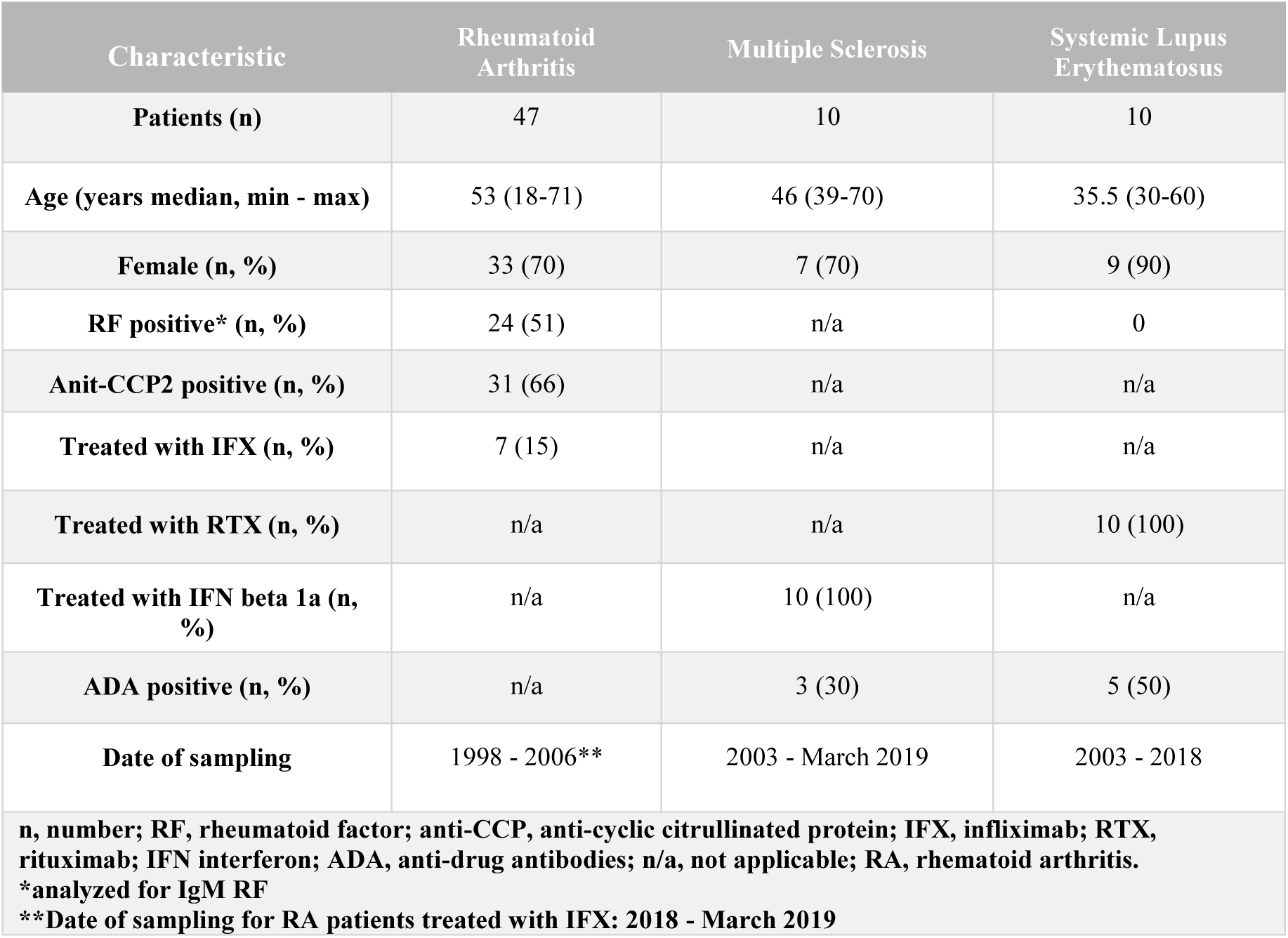
Patients’ characteristics.

## Methods

### Rheumatoid factor detection

Analysis of RF of the IgA, IgG and IgM isotypes of EIRA and SLE samples was performed using the EliA immunoassay on the Phadia 2500 instrument and the cutoff values as stated in the manufacturer’s instructions (Phadia GmbH, Uppsala, Sweden).^19^ Serum samples of RA patients treated with IFX were analyzed for IgM RF using laser nephelometry technique.

### Serological detection methods

A total of 19 commercial serological assays were evaluated in this study and compared to an in-house assay. Two ELISA and 17 rapid diagnostic lateral flow assays (LFA) were tested. These tests were assigned a letter from A – S (Table 2) and referred to as such in text and figures in this study. The brand name, antigen, manufacturer determined specificity and sensitivity, are outlined in Table 2. All tests were performed according to manufacturer instructions and using serum.

**Table 2.** Description of the SARS-CoV-2 serological assays and the test codes used in this study.

| Test Code | Manufacturer | Kit Name | Antigen/Target | Catalogue number | Company reported assay specificity |
| --- | --- | --- | --- | --- | --- |
| **A** | Zhuhai Livzon Diagnostics Inc. (China) | COVID-19 IgG/IgM Lateral flow Rapid Test Cassette | Nucleocapsid protein | Not specified | IgM: 99.7%<br>IgG: 99.4% |
| **B** | Epitope Diagnostics, Inc., San Diego, USA | EDI Novel Coronavirus COVID-19 IgG Elisa Kit | Nucleocapsid protein | KT-1032 | IgG: 100% |
| **C** | Jiangsu Medomics medical technology Co., Ltd, China | Rapid IgM-IgG combined Antibody Test Kit for SARS-CoV-2 (ICA | Spike protein (RBD MK201027) | 201030 | Not specified |
| **D** | Salafa Oy, Salo, Finland | Salacor (Biohit) SARS-CoV-2 IgG/IgM rapid test kit | Nucleocapsid protein | COV-01-S | IgM: 99.2%<br>IgG: 99.9% |
| **E** | Salafa Oy, Salo, Finland | Sienna SARS-CoV-2 IgG/IgM rapid test kit | Spike protein (RBD) | 102222 | IgM: 100%<br>IgG: 98.8% |
| **F** | Liming Bio-Products Co.,Ltd. Jiangsu, China | StrongStep_SARS-CoV-2 IgM/IgG_REF502090_Antibody Rapid Test | Nucleocapsid and Spike protein | 502090 | IgM: 100%<br>IgG:98.7% |
| **G** | Zhejiang Orient Gene Biotech Co., Ltd. (China) | COVID-19 IgG/IgM Rapid Test Cassette alt. HEALGEN_ COVID-19 IgG/IgM Rapid Test Cassette (Whole Blood/Serum/Plasma_REF GCCOV-402a | Nucleocapsid and Spike protein <sup>37</sup> | GCCOV-402a | IgM: 98.46%<br>IgG: 98.46% |
| **H** | InTec Products inc., Haicang Xiamen, China | INTEC_ Colloidal Gold (whole blood/Serum/Plasma) Rapid SARS-CoV-2 Antibody (IgM/IgG) | Nucleocapsid protein <sup>37</sup> | ITP16001-TC25 | Combined IgM+IgG: 98% |
| **I** | Sugentech Inc., South Korea * | SGTi-flex COVID-19 IgM/IgG | Nucleocapsid and Spike protein<sup>1</sup> | COVT025 E | IgM: 98.3% (90% FDA August 2020)<br>IgG:100% |
| **J** | Xiamen Biotime Biotechnology Co., Ltd. China | SARS-CoV2 IgG/IgM Rapid Qualitative Test | Spike protein<sup>1</sup> | BT1301 | Not specified |
| **K** | Microgen Diagnostik GmbH, Germany * | recomWell SARS-CoV-2 IgG | Nucleocapsid protein | 7304 | IgG: 98.7% |
| **L** | Abbott Point of Care Inc. USA * | Panbio COVID-19 IgG/IgM Rapid Test Device | Nucleocapsid protein <sup>38</sup> | ICO -T402 | IgM:92.8%<br>IgG: 92.8% |
| **M** | SureScreen Diagnostics Ltd, UK | SureScreen Diagnostics COVID-19 IgG/IgM Rapid Test Cassette (Whole blood/serum/ Plasma) | Spike protein /RBD | COVID19 C | IgM: 99.2%<br>IgG: 99.2% |
| **N** | Wuhan Easy Diagnosis Biomedicine Co.,Ltd (China) * | COVID-19 (SARS-CoV-2) IgM/IgG Antibody Test kit | Not specified | SA-2-D | IgM: 100%<br>IgG: 100% |
| **O** | Zhuhai Encode Medical Engineering Co.,Ltd.,China * | SARS-CoV-2 IgG/IgM Rapid test | S1-RBD and nucleocapsid protein | RCD-422 | IgM: 100%<br>IgG: 100% |
| **P** | Jiangsu SuperbioBiomedical Co., Ltd, China | SARS-CoV-2 (COVID-19) IgM/IgG Antibody Fast Detection Kit (Colloidal Gold) | Spike and nucleocapsid protein | B00502 | IgG: 95.8%<br>IgM: 95.8% |
| **Q** | Lumigenex (Suzhou) Co., Ltd. China | Lumigenex SARS-CoV-2 IgG/IgM Antibody Rapid Test Kit | Spike and nucleocapsid protein | Not specified | Not specified |
| **R** | Wondfo, Guangzhou, China | Wondfo Biotech SARS-CoV-2 Antibody Test | Not specified | W195 | Combined: IgM+IgG:99.57% |
| **S** | Innovita (Tangshan) Biological Technology Co Ltd (China) * | 2019-nCoV Ab Test (Colloidal Gold) | Recombinant antigen | Not specified | IgM: 100%<br>IgG: 100% |

<sup>1</sup>EUA Authorized Serology Test Performance | FDA
\*Stated in the instructions to have tested interference with RA and/or RF or other autoantibodies

The results were compared to an in-house multiplex bead-based and validated SARS-CoV-2 serological assay developed at SciLifeLab and KTH Royal Institute of Technology as previously described ^20^. In brief, IgG reactivity was analysed in a high-throughput and multiplex bead-based format utilizing 384-well plates and FlexMap3D instrumentations (Luminex Corp) for read-out (22). Reactivity against three different in-house produced viral protein variants was used to differentiate between positive and negative samples: Spike trimers comprising the prefusion-stabilized spike glycoprotein ectodomain^21^ (expressed in HEK and purified using a C-terminal Strep II tag), Spike S1 subunit (expressed in CHO and purified with HPC4 tag), and the Nucleocapsid protein (expressed in E. coli and purified using an N-terminal His-tag). The antigens were immobilized on magnetic colour coded beads (MagPlex, Luminex Corp) and plasma/serum IgG that bound to the antigens were detected by an R-phycoerythrin conjugated goat anti-hIgG (Invitrogen, H10104). Reactivity against at least two out of the three viral antigens included in the panel was required for positive read out. The cut-off for seropositivity was defined as signals above the mean +6 SD of the 12 negative controls included in each assay, The method utilizing the combination of the three antigens has been found to have 99.2% sensitivity (99.6%, 99.2%, 96.7%, respectively, for the three antigens individually) and 99.8% specificity (98.9%, 99.1%, 98.4%, respectively, for the three antigens individually) based on 243 positive control (defined as >16 days after onset or positive PCR) and 442 negative control (defined as collected 2019 and earlier) samples.

### Commercially available ELISA kits

The two included Enzyme-Linked Immunosorbent Assays (ELISAs) were performed according to the manufacturers’ instructions. The first was the EDI Novel Coronavirus COVID-19 IgG Elisa Kit (Epitope Diagnostics, Inc., San Diego, USA) to detect IgG (test B). This is an IVD and CE marked indirect ELISA with plates coated with peptides from the SARS-CoV-2 nucleocapsid antigen. Specificity of this assay was determined by the manufacturer using anti-influenza A, anti-influenza B, Hepatitis C virus (HCV), anti-nuclear antibodies (ANA) and respiratory syncytial virus (RSV). The cut-off for positivity was determined according to the manufacturer’s instructions. The manufacturer states that a positive result may be due to past or present infection with SARS-CoV-2 but not due to other coronavirus strains, such as coronavirus HKU1, NL63, OC43, or 229E.

The second ELISA used to detect IgG against SARS-CoV-2 was the recomWell SARS-CoV-2 IgG Elisa kit (Microgen Diagnostik GmbH, Germany (test K)). This assay is also an indirect ELISA which uses highly purified recombinant nucleocapsid protein from SARS-CoV-2 as an antigen. The manufacturer had determined the potential interference of antibodies against other pathogens that might induce clinical symptoms similar to those of a SARS-CoV-2 infection (including for example seasonal coronaviruses, influenza A virus, RSV, Mycoplasma pneumoniae, Chlamydia pneumoniae). In addition, they also tested specificity using samples from people with conditions that present with atypical immune system activity, including EBV infection, pregnancy, and ANA- and-RF-positive subjects. The cut-off for positivity was calculated according to the test instructions.

### Commercial Lateral Flow Assays

LFAs are designed to enable point of care analyses and can generate fast results with read-outs as bands in small cartridges. These rapid lateral flow tests are developed for whole blood, serum and plasma. At time of testing, the appropriate volume of serum was applied to the designated well and then the buffer was added. After the recommended incubation period, the presence and intensity of the bands were investigated and graded from negative to four levels of positivity by the operator.

### Statistical Analyses

Raw data were analysed as per individual commercial test instructions. Rate of false positive signals were determined as the number of positive samples divided by the total number of samples tested for each assay. Statistical analyses and figures were generated using GraphPad Prism (version 8.2.1). The statistical significance of having a reaction against RF compared to serum without RF in RA patients were calculated with Fishers exact test. The other groups were too small to make any meaningful statistical evaluations and thus these results are only presented as descriptive analyses.

## Results

### Serology Assay Specificities

#### Commercial LFA and ELISA assays

Serum samples from 47 RA patients (with two samples from one of the patients), 10 SLE and 10 MS patients were evaluated on 19 SARS-CoV-2 commercial serological assays and compared to an in-house developed multiplex bead-based assay.^20^ The overall results of all 68 samples are illustrated in figure 1A. A total of six commercial LFAs (test A, G, H, J, R and S) reached 100% specificity for both IgG and IgM including all chronic inflammatory disease cohorts’ patients (*n*=67). Notably, all samples from MS patients (*n*= 10) were negative for both IgM and IgG in all 20 assays.

**Figure 1.**
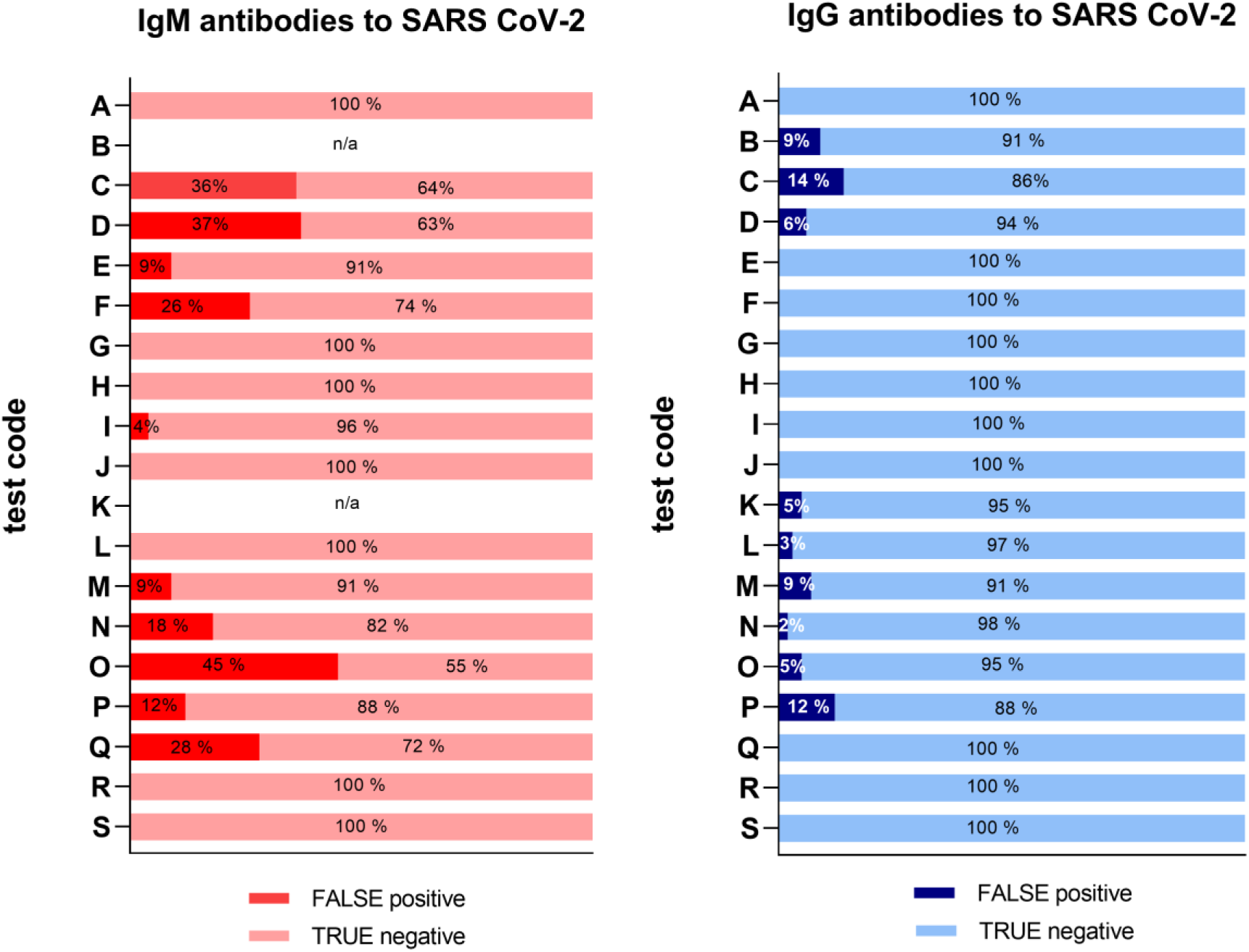
Overview of false positive results in all samples for 19 different serological tests. Six LFA tests (A, G, H, J, R & S) did not show any false positives at all. For the rest of the tests the false positivity rate ranged between 2% and 45%. The test code keys are described in Table 2. The two ELISA assays (test B and K) were only tested for IgG.

For the 17 LFAs evaluated for specificity using 25 RA samples (from 24 patients) which were positive for RF, 10 assays had unspecific signal detected for at least one immunoglobulin isotype (Figure 2). Five assays had unspecific signal for both IgM and IgG in a few to the majority of the samples (test C: IgM 19/20, IgG 8/20; test D: IgM 19/20, IgG 2/20; test M: IgM 4/20, IgG 3/ 20; test N: IgM 6/20, IgG 1/20; and test P: IgM 1/20, IgG 1/20). Unspecific IgM signal, without unspecific IgG signal, was detected in four LFAs (test E: 5/20; test F: 16/20; test O: 20/20; and test Q: 19/20). In one LFA, only the IgG test gave unspecific signal (test L: 1/20). In contrast, only five assays detected unspecific signal in naive RA samples that were RF negative (*n*= 23), with five detecting IgM and one detecting IgG (test D: IgM 1/23; test F: IgM 1/23; test M: IgM 2/23, IgG 2/23; for test N: IgM 1/23; for test O: IgM 1/23) (Figure 2). None of the two ELISAs (test B and K) gave any false positive signals with these samples. Due to insufficient sample volume, these ELISA tests could not be verified as extensively as the other tests (supplementary table 1).

**Figure 2.**
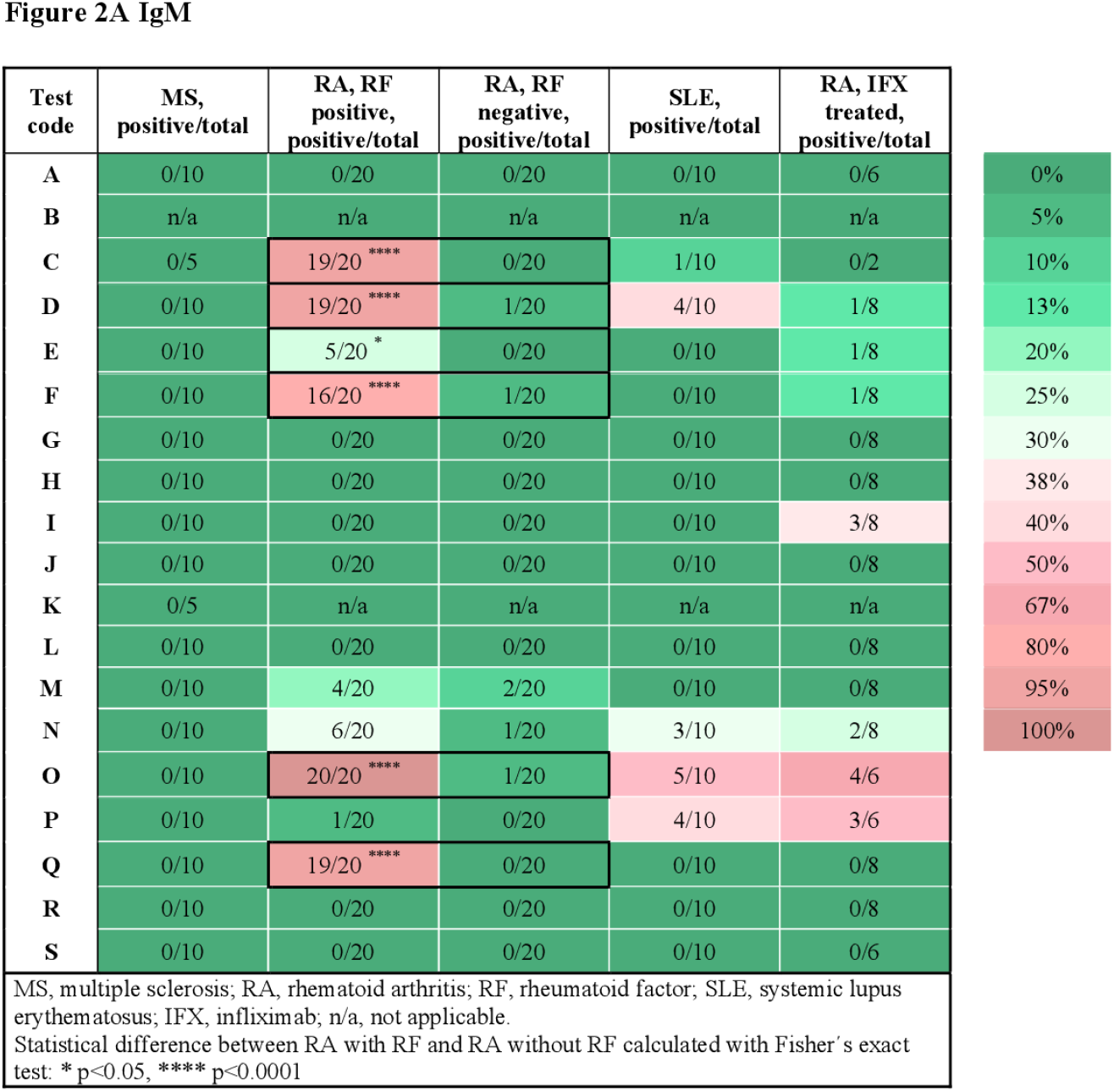
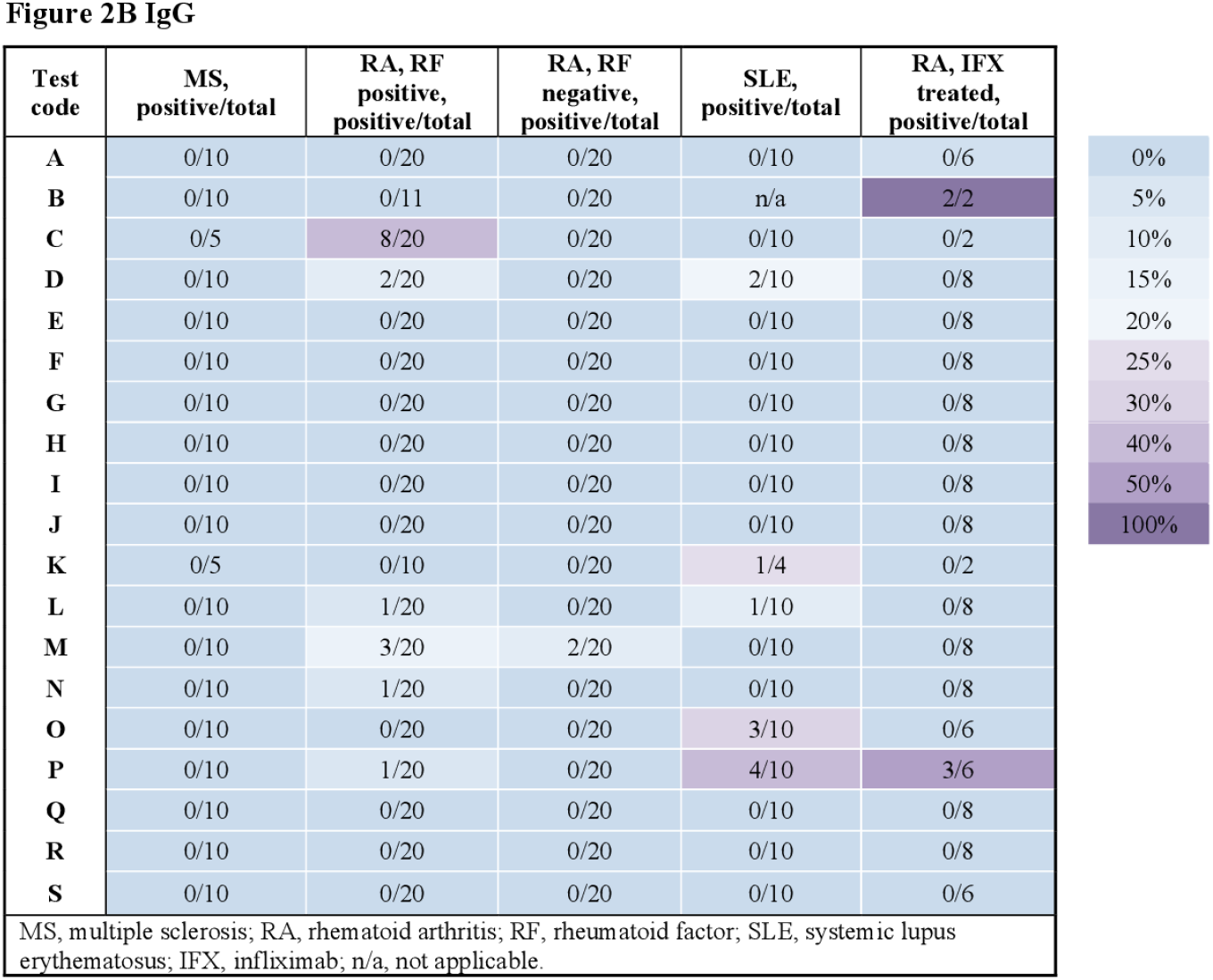
Percentage of the false positive test results for A) IgM and B) IgG with samples from MS patients, DMARD naïve RF positive RA patients (n= 20), RF negative (n=20) RA patients, SLE patients (n=10), infliximab treated RA patients (n=8). The test code keys are described in Table 2. Stars indicate significant difference between RF status in RA patients, using Fisher exact test, * p<0.05, *** p<0.0001.

When using IFX treated-RA patients as negative controls (patients *n*=7, samples *n*=8) (Figure 2), unspecific signal was detected for IgM in seven assays (test F: 1/8; test D: 1/8; test E: 1/8; test I: 3/8; test N: 2/8; test O: 4/6; and test P: 3/6) and for IgG in two assays (test B: borderline positive signal in 2/2 samples and test P: 3 of 6 samples). Two samples were from one individual at two time points; prior to second infliximab infusion and after 9 months on treatment initiation. These two samples (IFX1 and IFX2 in supplementary table) were both borderline positive in test B, and the sample taken after 9 months (IFX1) was positive in test E for IgM only.

### Serology Assay Specificities compared to occurrence of RF isotypes

The levels of IgG, IgM and IgA RF were very high in the RF positive RA samples (n=20), as these had been selected as such. Thus, associations between specific RF isotypes and false positive IgM/ IgG anti-SARS-CoV-2 response could not be analyzed. The SLE samples (n=10), on the other hand, had a diversity of RF isotypes, i.e. none of the patients had IgM RF, three had IgA RF and one was positive for IgG RF. No associations were identified between RF isotypes and false positive anti-SARS-CoV-2 IgM or IgG signal in the SLE samples. However, there was a trend towards higher titers of IgA RF and absence of false positive IgM/ IgG anti-SARS-CoV-2, i.e. two samples with high IgA RF levels (SLE2 with 551 IU/ml and SLE 7 with 26 IU/ml respectively in supplementary table 1) were negative in all tests; and two samples (SLE1 and SLE8 in supplementary table 1) with the highest number of tests with false positive signals were negative for IgA RF. We also found that one RF negative SLE sample was IgM positive in two tests (C and N) and another RF negative SLE sample was both IgM and IgG positive in two tests (O and P). No associations were identified between anti-CCP2 antibodies or C1q-binding immune complexes and false positive IgM/IgG anti-SARS-CoV-2 response.

#### SciLifeLab and KTH in-house validated SARS-CoV-2 serological assay

Due to insufficient sample volume only 66 of the 68 samples were analysed using the in-house developed multiplex bead-based assay for IgG detection as described above.^20^ All samples analysed using this method were classified as negative. The only two samples not included were the two infliximab treated samples from the same patient (supplementary table 1).

## Discussion

Serological assays are necessary tools in a pandemic, both for determining the proportion of the population already subjected to the infection and for the individual to confirm past infection and present immunity. In the case of SARS-CoV-2, it seems that a proportion of the individuals who have been infected do not develop antibodies, at least not as determined by currently available serological assays ^22^. There also seem to be some pre-existing immunity present in the population, as determined by memory T cell reactivity^23 24^ and the estimated prevalence of infected individuals in comparisons to the proportion that succumb in severe disease.^25^

To elucidate these issues, we have to rely on the serological assays. Therefore an independent verification of sensitivity and specificity of such assays is often required. The specificity of an assay is defined as the ability to correctly assign negative samples as negative. It is determined by a selection of samples that are supposed to be negative for the new infection and typically the negativity is guaranteed by having samples collected before SARS-CoV-2 emerged. When serological assays against viral antigen are developed, one major concern is regarding the cross-reactivity against similar viruses ^15^. SARS-CoV-2 serological assays using antigens that cross-react with antibodies generated towards other coronaviruses will not be approved, since they would not serve the purpose of answering the clinically and epidemiologically important questions of who has developed antibodies against the new virus.

The aspect of immunity against SARS-CoV-2 is of particular importance to persons with chronic inflammatory diseases, given the concerns that treatments or their underlying disease might render them less able to fight the infection, establish immunity or respond to vaccinations. Only a few viral serology assays on the market will have tested for interferences using serum from patients with chronic inflammatory diseases, primarily because these are typically not easily accessible for manufacturers. However, these types of sera are notorious for interfering in immunological assays, giving higher background and unspecific signals. For instance, in the drug immunogenicity field, when validating assays for determining ADA, it is recommended to account for such unspecific signal during assay development and validation. This is achieved using a cohort of baseline samples in clinical trials from the targeted patient population who are treatment naive to the biological drug for which the ADA assay is developed.^16 17^ For serological assays used to detect viral infections, such interference might not be discovered until you start to do extensive screening of larger populations. This would become particularly notable, and give a false impression of exposure and immunity, if there is an interference by serum factors from patients with common diseases that have a frequency in the same magnitude as the studied infection. These serum factors could include autoantibodies, biological drugs, ADA, or aggregates and immune complexes formed by one or several of these components together. To complicate the matter further, there are indications of that SARS-CoV-2 infection and COVID-19 disease might trigger these autoantibodies.^26^

In the present study, a selection of samples from patients with chronic inflammatory diseases were used to determine the specificity of a range of SARS-CoV-2 serological assays. We found that false positive results occur in the majority of the serological assays evaluated (Figure 1). Most notably, samples from RA patients with high levels of RF resulted in a false positive signals in several assays (Figure 2A and B). As RF binds to the constant parts of IgG, this could precipitate other antibodies present in an immunoassay in an unspecific way. These unspecific positive signals might not only give false indication of protective immunity to SARS-CoV-2 for an individual, but might also give an incorrect picture of the proportion of the population exposed to the infection during larger screenings. Most of the false positive signals where detected in the IgM assays, as has been noted by others,^15^ which might be in line with the broader low affinity quality of the IgM antibodies, as compared to IgG class switched and affinity matured antibodies. Other studies have reported about this issue with different interpretations. One study using only one test (Innovita Biotechnology Co, Tangshan, China) reported that there was no interference with serum from persons with autoimmune disease,^27^ which we can confirm here for the Innovita LFA (test S).

Serum from patients with SLE have a high abundance of autoantibodies, which are predominantly directed against double-stranded DNA. However, many other targets have also been described and the isotypes and specificities of these autoantibodies correlates with the symptoms of the disease ^28^. Although SLE is a less prevalent disease than RA, serum samples from these patients contributed essentially to the false positive signals in the present study.

Rheumatoid factor was the first autoantibody discovered in RA. According to different studies, RF has limited specificity for RA (from 48% to 92%),^29^ since it can also be present in healthy controls and patients with other autoimmune and non-autoimmune diseases, such as chronic infections and cancer, and now also in COVID-19 survivors.^26 29-32^ Since RF are heterophilic and can involve different immunoglobulin classes (IgM, IgG and IgA), we characterized these further. IgM-RF is the isotype commonly measured in most clinical laboratories, and detected in 60-80% of RA patients,^29 32^ but might appear also in other diseases.^29 32 33^ In the current study, we were not able to detect any specific associations between occurrence of RF IgM or RF IgG and false positivity for IgM/ IgG anti-SARS-Cov-2 in RA patients, since the RF positive RA sera were specifically selected to be highly positive for all RF isotypes simultaneously. Regarding the SLE samples, no positive associations were identified between specific RF isotypes and false positive signal. However, there was a trend towards higher titers of IgA RF and absence of false positive IgM/ IgG anti-SARS-Cov-2. The false positive signals in SLE samples observed in the present study might be explained by other autoantibodies such as ANA, anti-Sm/RNP, anti-Ro/La, anti-dsDNA etc. The exact biochemical interactions with RF in the SARS-CoV-2 serological assays have to be investigated further.

It could be argued that the unspecific signals detected in this study might actually be due to some underlying immunity, if there are mechanisms such as molecular mimicry behind the triggering of autoimmunity^34 35^ and these would also, hypothetically, work in the reverse direction. However, a more plausible explanation is that it is due to a technical difficulty in the assay development and thus one should not assume that these signals confirm any immunity against infection.

It should also be noted that samples used to determine the specificity of SARS-CoV-2 serological assays will be highly variable between manufacturers and often not reported in detail in the assay labels or information inserts. However, some manufacturers and vendors are aware of this issue and have included this in the information about the assay. Encode (test O) for example, report that specimen containing higher titers of heterophobic antibodies or rheumatoid factors may affect the results. With the RF positive RA samples used in the present study, 100% reacted in the IgM assay and 5% in the IgG assay, but also both RF positive and RF negative SLE samples gave signal. The Easy Diagnosis (test N) have tested RF, human anti mouse antibody (HAMA), and anti-nuclear antibody (ANA), and claim that they do not interfere with the kit, but in the present study we show that 5-30% of the samples gave false positive signal. Innovita (test S) reported that samples positive for RF, ANA, HAMA, SLE have been analyzed, and they do not report cross reactivity in their test, as could also be independently verified by us. Microgen Diagnostik (test K) reported cross-reactivity with RF, but we could only identify cross-reactivity in the IgG assay with an RF negative SLE sample. Abbott (test L) reported that they tested samples positive for RF (3/3), HAMA (3/3), and ANA (3/3), and that these did not affect the performance of the test. However, we see two false positive signals in the IgG test for RF positive RA and SLE samples. Sugentech (test I) reports no cross-reactivity with anti-human IgG, IgM, IgA and IgE, but here we report false positive signal in three samples from an IFX treated patients. Notably, for Sienna (test E), of the two samples tested from an IFX treated patient, one sample taken before and one sample taken after infliximab treatment, only the one on treatment gave false positive signal in the IgM test. This rises additional concern, given how many people who currently are treated with infliximab. However, since none of the other samples from IFX treated RA reacted, it speaks against it. The second sample from the IFX RA had high ADA, which then potentially could be the interfering factor. These findings have to be verified in a larger cohort. The Sienna (test E) also gave false positive signal for five of the RF positive untreated RA samples and since the IFX treated RA also was positive for RF, this might also be an interfering factor.

It should also be noted that several serology tests did not show any false positive results with these complicated sera and thus there are methods to avoid unwanted interference. A suggested method to resolve the RF issue, at least in ELISA tests, is to use urea for dissociation of the interfering signals.^36^ There are some limitations in this study. Firstly, there are over 200 SARS-CoV-2 serology assays available on the market and with the limited set of stored samples, we could only analyse a fraction of these. For reliable use of serological assays for patients with chronic inflammatory diseases, each assay would need to be individually analysed with negative serum from that patient population, before one starts to screen that patient group. Here we can only report the specificity in relation to MS, RA and SLE. Due to the limited availability of sample material, only one test result per sample, per assay, was retrieved and it was not possible to further elucidate the molecular mechanism behind the positive signals.

Secondly, the LFA’s are primary made for whole blood, to enable individual to do a rapid test with a drop a blood from the fingertip, but here we only had stored serum to use for testing. However, all of the assay also indicate that they work with serum and plasma. Given that the serum from MS patients did not give any signal in any assay, the false positive signals detected in this study is most probably not an issue of having a different matrix, but more likely the unspecific antibody contents of the serum.

In conclusion, serological assays are sensitive to interfering antibodies, especially from persons with autoimmune diseases. There is a trade-off between requiring extensive screening for unspecific binding in these assays and the harm the delay the process of making these assays available for mass screening might cause, so a cost benefit analysis has to be made on both national and global level. However, if persons with autoimmune disease, health care providers and decision makers are aware about this issue, they could adapt the testing strategy and interpretation of the results accordingly. To enable such informed decisions, it would be helpful if information about which types of samples have been used for validation of specificity is stated in the label of all of the tests.

## Data Availability

The raw data supporting the conclusions of this manuscript will be made available by the authors, without undue reservation, to any qualified researcher.

## Author Contributions

All authors contributed to conceptualization, execution, writing, review, and editing of the manuscript. All authors approved the final version of the manuscript.

## Ethics

Ethics Review Authority in Stockholm and Gothenburg (2020–23/04, dnr 2020-01649, 2012/1550-31/3, dnr 96-174).

## Acknowledgements

We would like to thank a number of people who have contributed to this study in many ways including technical, practical and financial support as well as valuable intellectual discussions. These are Anna Mattson, the Crowdfighters volunteers, Lena Israelsson, Monica Hansson, Lars Klareskog, Lars Alfredsson, Francesca Carlisle, Simona Marzorati and Luciana Mazzocchi, Wefightcovid volunteers, Eva Sundman (Head of Medical and Scientific Director REMEO), Anna Cunningham at SLSO, Peder Olofsson, Erik Eberhardsson, Martin Schalling, Joakim Dillner and colleagues, Jan Hillert, Linda Hahn, Anders Struknäs (Feronia Nordic AS), Richard Grönevall, Karina Persson, Simon Strandh at Carmona, Cecilia Hellström, Eni Andersson, Jennie Olofsson at SciLifeLab and KTH, Ebba Carbonnier and Peter Nordström at Swelife/Vinnova, Anna Ridderstad-Wollberg and Anders Kallin at RISE.

