## Supplementary figures and images for "SARS-CoV-2 serological tests can generate false positive results for samples from patients with chronic inflammatory diseases"

### Supplemental Table 1

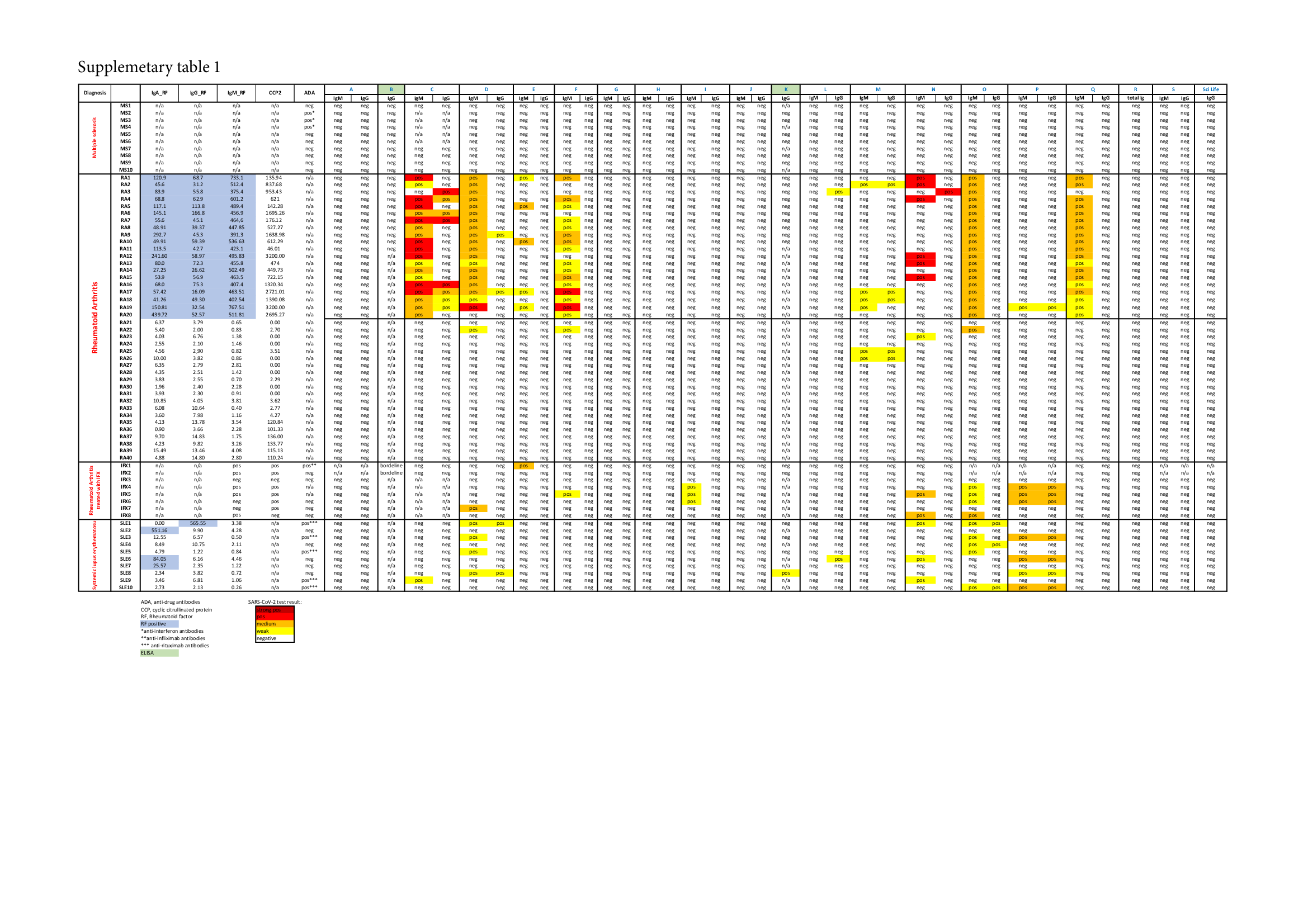
